# Association of anti-Ro-52 positivity with cardiovascular outcomes in patients with anti-synthetase syndrome

**DOI:** 10.64898/2026.07.04.26357290

**Authors:** A.V. Potharazu, J.H. Chung, L. Yanek, W. Kelly, N. Gilotra, L. Adamo, J. Paik

## Abstract

**Background:** Anti-synthetase syndrome (ASyS) is a subgroup of idiopathic inflammatory myopathies that is increasingly recognized as a distinct entity with features of myositis, interstitial lung disease, inflammatory arthritis, and Raynaud’s phenomenon. Co-reactivity with anti-Ro-52, an antibody directed against the Ro-52 E3 ubiquitin ligase, has been shown to be associated with progressive interstitial lung disease within this patient population. However, less is known regarding the association of anti-Ro-52 positivity with cardiovascular outcomes.

**Methods:** A sub-cohort of patients with anti-synthetase antibodies at a large single institution center was retrospectively analyzed to define presence of anti-Ro-52 positivity (defined as anti-Ro-52 titer greater than or equal to 11 utilizing the line immunoblot platform, Euroline Autoimmune Inflammatory Myopathies, EuroImmun Diagnostics, Lubeck, Germany). Patients who did not meet 2017 ACR/EULAR classification criteria for idiopathic inflammatory myopathies were excluded from the final analysis. Cardiovascular outcomes ascertained via retrospective chart review included atrial fibrillation, left bundle branch block, right bundle branch block, pulmonary hypertension (confirmed via right heart catheterization), heart failure with reduced ejection fraction (HFrEF, defined as ejection fraction less than or equal to 40%), acute coronary syndrome (based on clinical diagnosis and angiography if available), and myocarditis (based on clinician diagnosis and either cardiac MRI or troponin elevation). When a pre-specified cardiac outcome was identified, the date of onset was recorded. Differences in proportions were analyzed via Chi-squared and Fisher’s exact tests, and time-to-event analyses were performed via Cox Proportional Hazards Models, incorporating a false discovery rate correction for multiple outcomes. All analyses were performed using SAS v9.4.

**Results:** 88 patients were included in the final analysis, of whom 69 (78.4%) were categorized as anti-Ro-52 positive. Patients with anti-Ro-52 positivity had a higher maximum recorded serum creatine kinase (median 1297 vs 395 units per liter, p = 0.042). No significant associations between anti-Ro-52 positivity and the pre-defined cardiovascular outcomes were found over median follow up time of 12.5 years.

**Conclusions:** In a large, single-center cohort of patients with ASyS, anti-Ro-52 positivity was not associated with an increased burden of negative cardiovascular outcomes, including the onset of pulmonary hypertension. Future studies may seek to further elucidate the mechanisms underlying the pleiotropic effects of anti-Ro-52 antibodies on the cardiopulmonary system.

## Introduction

Idiopathic inflammatory myopathies constitute a clinically heterogeneous group of systemic auto-immune disorders with a shared pathophysiology of immune-mediated muscle injury.^1^ Advances in serologic phenotyping through both myositis-specific and myositis-associated autoantibody profiling have driven the identification of distinct subgroups of inflammatory myopathies, with the potential to improve precision in diagnosis, prognostication, and treatment.^2–4^ Anti-synthetase syndrome is a rare but increasingly recognized form of inflammatory myopathy defined by the presence of one or more anti-aminoacyl tRNA synthetase autoantibodies.^1^

Interstitial lung disease is a major complication of anti-synthetase syndrome and is associated with increased morbidity and mortality.^5,6^ Of the myositis-associated autoantibodies, anti-Ro-52, which targets the E3 ubiquitin ligase Ro-52, has emerged as a major independent predictor of the development of interstitial lung disease across a variety of connective tissue diseases, including anti-synthetase syndrome.^7^ It is assumed that mortality from interstitial lung disease is in part mediated through downstream associated secondary cardiovascular outcomes, including the onset of group I and group III pulmonary hypertension and right ventricular dysfunction.

Cardiac involvement in inflammatory myopathies and anti-synthetase syndrome spans a broad spectrum beyond pulmonary hypertension, including conduction abnormalities, coronary artery disease, cardiomyopathy, and myocarditis. Cardiac comorbidities in anti-synthetase syndrome have heterogenous presentations with variability in timing of onset and severity and are associated with adverse prognosis.^8,9^

Ro-52, also referred to as Tripartite Motif Containing 21 (TRIM21), is a key mediator of immune homeostasis in both type I interferon and interleukin-17-mediated pathways, playing both pro-inflammatory and anti-inflammatory roles.^10^ With regards to cardiomyocytes, Ro-52 has been shown to mediate pathologic myocardial injury in mouse models, with Ro-52-knockout mice having attenuated pathologic remodeling.^11,12^ Ro-52 has also been recently shown to mediate pathologic remodeling after myocarditis.^13^

To our knowledge, no study has directly examined the association between anti-Ro-52 positivity and cardiac comorbidities in patients with anti-synthetase syndrome. In this study, we leveraged a well-characterized, large single center cohort to evaluate whether anti-Ro-52 positivity was associated with the frequency and time-to-onset of a pre-specified set of cardiovascular outcomes, including atrial fibrillation, conduction abnormalities, pulmonary hypertension, heart failure with reduced ejection fraction, acute coronary syndrome, and myocarditis. Given the well-described association between anti-Ro-52 autoantibodies and interstitial lung disease, we hypothesized that anti-Ro-52 positivity would be associated with an increased burden of cardiac disease.

## Methods

### Sub-Cohort of Patients with Anti-Synthetase Syndrome

A sub-cohort of patients with anti-synthetase syndrome enrolled in the Johns Hopkins Myositis Center was retrospectively queried from 2003 to 2025. Anti-synthetase syndrome was defined as patients meeting the 2017 ACR/EULAR criteria for idiopathic inflammatory myopathy and the presence of an anti-synthetase antibody (anti-Jo1, anti-PL12, anti-PL7, anti-EJ, and/or anti-OJ). To maintain focus on anti-synthetase syndrome, patients with other forms of idiopathic inflammatory myopathy, such as immune-mediated necrotizing myopathy, inclusion body myositis, and overlap myositis with systemic lupus erythematosus or systemic sclerosis were excluded.

Myositis specific antibodies were assayed on banked sera using a commercially available line immunoblot platform (Myositis profile, Euroimmun Diagnostics, Lübeck, Germany). A positive result was identified as a value of 11 or greater as recommended by the manufacturer.^14^

Patient charts were manually reviewed for the presence of atrial fibrillation (based on a cardiologist-confirmed EKG read), left bundle branch block, right bundle branch block, pulmonary hypertension, heart failure with reduced ejection fraction (defined as left ventricular ejection fraction ≤ 40%), acute coronary syndrome (defined by clinician diagnosis), history of coronary revascularization, and myocarditis (defined by clinician diagnosis and either troponin elevation or consistent cardiac MRI findings) through 16 April 2025 (Supplement 1) or until the recorded date of death, whichever came first. When noted, the earliest date of onset of the comorbidity was recorded. Patients with pulmonary hypertension were defined as having echocardiographic findings with an estimated right ventricular systolic pressure ≥ 35 mmHg. A subgroup analysis was performed for patients with pre-capillary pulmonary hypertension confirmed by invasive hemodynamic analysis, which was defined as documented findings of a mean pulmonary artery pressure greater than 20 mmHg, a pulmonary arterial wedge pressure ≤15 mmHg, and a pulmonary vascular resistance greater than 2 Wood units.^15^ Patients with presumptive pulmonary hypertension via transthoracic echocardiography but without invasive hemodynamic data for confirmation were included in the overall pulmonary hypertension analysis but excluded from the subgroup analysis for patients with invasive hemodynamic data. For heart failure with reduced ejection fraction, the nadir ejection fraction was recorded.

### Statistical Analysis

Data were presented as frequencies and proportions or means with standard deviations or medians with 25^th^ and 75^th^ percentiles. Associations between Ro-52 positivity and demographic and clinical variables, and clinical outcomes of interest (atrial fibrillation, left bundle branch block, right bundle branch block, pulmonary hypertension, heart failure with reduced ejection fraction, acute coronary syndrome, history of coronary revascularization, and myocarditis) were assessed using Chi-squared or Fisher’s exact tests, and presented as odds ratios with 95% confidence intervals. Associations between Ro-52 positivity and continuous variables of interest were assessed using t-tests or Wilcoxon tests. Associations between Ro-52 positivity and outcomes with a minimum of five events (atrial fibrillation, right bundle branch block, pulmonary hypertension, heart failure with reduced ejection fraction, history of coronary revascularization, myocarditis) were assessed with Cox proportional hazards regression models, predicting time-to-event as time from symptom onset of anti-synthetase syndrome to event, with censoring at death or last follow-up at 4/16/2025. Time to events was visualized with Kaplan-Meier curves and compared with log-rank tests. The Benjamini-Hochberg false discovery rate was used to determine statistical significance accounting for multiple comparisons. Analyses were performed using SAS v9.4 (SAS Institute Inc., Cary, NC).

## Results

111 patients were initially identified as having anti-synthetase antibodies, of whom 70 [63.1%] were anti-Jo1 positive. After removing patients who did not meet 2017 ACR/EULAR classification criteria for idiopathic inflammatory myopathies, a total of 88 patients were included in the final analysis. All patients who had anti-Jo-1 positive antibodies were included in the final analysis. 14 patients with anti-PL12 antibodies, 6 patients with anti-PL7 antibodies, 2 patients with anti-EJ antibodies, and 1 patient with anti-OJ antibodies were removed.

69 patients were categorized as anti-Ro-52 positive, and 19 patients were categorized as anti-Ro-52 negative. Patients with anti-Ro-52 positivity had a higher maximum recorded serum creatine kinase (median 1297 versus 395 units per liter, p = 0.0415) and a trend towards a younger age of onset (mean 44.8 ± 11.2 years versus 50.3 ± 11.0 years, p = 0.058). Although patients with anti-Ro-52 positivity had a higher frequency of ILD diagnosed via imaging criteria on high resolution computed chest tomography, the difference did not reach statistical significance when comparing patients with available data (frequency 46 [92.0%] versus 8 [72.7%] with data not available for 27 patients, p = 0.10).

Frequencies of cardiac outcomes with associated odds ratios are presented for the pre-specified outcomes with a p-value corrected based on false discovery rate in the right-most column (Table 2). Except in three instances, all outcomes occurred after the onset of symptoms from anti-synthetase syndrome. Anti-Ro-52 positivity was not associated with a difference in frequency of atrial fibrillation, left bundle branch block, pre-capillary pulmonary hypertension, heart failure with reduced ejection fraction, coronary revascularization, or myocarditis. Although initial analysis suggested an inverse association between anti-Ro-52 positivity and frequency of acute coronary syndrome, this difference was not found to be statistically significant after false discovery rate correction.

**Table 1.**
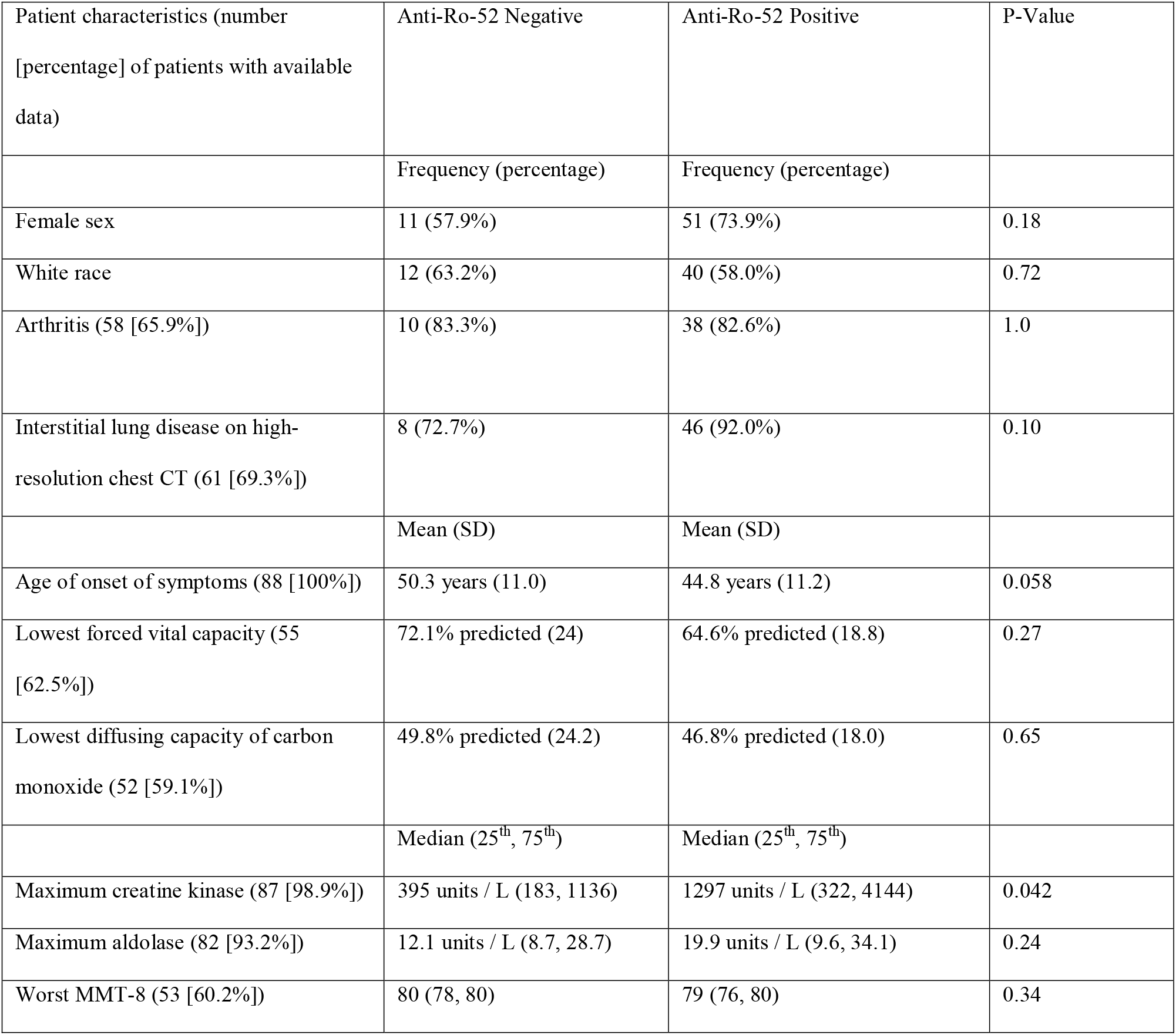
Baseline demographic variables stratified by anti-Ro-52 positivity. Frequency of patients with available data noted for each demographic variable due to missing data.

**Table 2.** Frequency of cardiac outcomes stratified by Ro-52 positivity.

| Outcome | Anti-Ro-52 Negative<br>(frequency [percentage]) | Anti-Ro-52 Positive<br>(frequency [percentage]) | Odds Ratio (95% Confidence Interval) | Unadjusted P-Value | P-Value After False Discovery Rate Correction |
| --- | --- | --- | --- | --- | --- |
| Atrial fibrillation | 3 (15.8%) | 10 (14.5%) | 0.90 (0.22 - 3.68) | 1.0 | 1.0 |
| Left bundle branch block | 0 (0%) | 1 (1.45%) | --- | 1.0 | 1.0 |
| Right bundle branch block | 2 (10.5%) | 4 (5.80%) | 0.52 (0.09 - 3.10) | 0.61 | 1.0 |
| Pulmonary hypertension via echocardiography | 6 (31.6%) | 21 (30.4%) | 0.95 (0.32 - 2.83) | 0.92 | 1.0 |
| Pulmonary hypertension confirmed by invasive hemodynamics (n=82) | 5 (27.8%) | 16 (25.0%) | 0.87 (0.27 - 2.81) | 0.81 | 1.0 |
| Heart failure with reduced ejection fraction (LVEF ≤ 40%) | 4 (21.1%) | 10 (14.5%) | 0.64 (0.17 - 2.31) | 0.49 | 1.0 |
| Acute coronary syndrome | 2 (10.5%) | 0 (0%) | --- | 0.04 | 0.40 |
| Any coronary revascularization | 1 (5.2%) | 2 (2.9%) | 0.54 (0.05 - 6.27) | 0.52 | 1.0 |
| Myocarditis | 1 (5.3%) | 3 (4.4%) | 0.82 (0.08 - 8.3) | 1.0 | 1.0 |

No significant differences in time of onset were noted for atrial fibrillation, right bundle branch block, pulmonary hypertension, heart failure with reduced ejection fraction, or myocarditis (Table 3, Figure 1). The hazard ratio for time to event was ≤1 for all cardiovascular outcomes, but all 95% confidence intervals were not found to be statistically significant. This finding applied as well to log-rank tests on Kaplan-Meier curves (Supplement 2).

**Table 3.** Time-to-event analysis via Cox Proportional Hazards Models of cardiac comorbidities in patients with anti-synthetase syndrome, stratified by Anti-Ro-52 positivity. Three cases were dropped due to outcome presence at the time of ASyS symptom onset (n=1 for atrial fibrillation, n=1 for pulmonary hypertension, and n=1 for HFrEF).

| Outcome | Hazard Ratio (95% Confidence Interval) | P-Value | P-Value After False Discovery Rate |
| --- | --- | --- | --- |
| Atrial fibrillation | 0.87 (0.24 - 3.18) | 0.84 | 0.84 |
| Right bundle branch block | 0.49 (0.09 - 2.71) | 0.42 | 0.84 |
| Pulmonary hypertension via echocardiography | 0.87 (0.35 - 2.2) | 0.77 | 0.84 |
| Pulmonary hypertension confirmed by invasive hemodynamics ($n = 100$) | 0.88 (0.32 - 2.44) | 0.47 | 0.84 |
| Heart failure with reduced ejection fraction | 0.65 (0.20 - 2.09) | 0.81 | 0.84 |

## Discussion

In this study, we sought to evaluate the association of anti-Ro-52 positivity with cardiac comorbidities and pulmonary hypertension in patients with anti-synthetase syndrome. No significant differences in frequency or time to onset of atrial fibrillation or bundle branch blocks, cardiomyopathies, or acute coronary syndrome were found. In addition, we did not find a significant difference in frequency or time to onset of pulmonary hypertension despite previous studies describing an association with interstitial lung disease in patients with anti-Ro-52 positivity.^16^ These highlight a possible unexpected disparate prognostic significance of anti-Ro52 positivity in terms of risk of pulmonary versus cardiac comorbidities in this patient population that requires further elucidation. However, the strength of the conclusion may be limited by the sample size and characteristics of this study’s cohort.

Advances in classification of inflammatory myopathies, including differentiation of anti-synthetase syndrome, have been driven by improvements in serological subtyping. Accordingly, auto-antibody profiling is playing a larger role in the phenotyping of inflammatory myopathies. Ro-52, an E3 ubiquitin ligase, is upregulated in pro-inflammatory environments and plays a role in the pathogenesis of autoimmunity.^17^ In addition to idiopathic inflammatory myopathies, anti-Ro-52 antibodies are found in patients with systemic lupus erythematosus, systemic sclerosis, Sjogren’s syndrome, and auto-immune hepatitis.^18^ Anti-Ro-52 positivity has a well-described association with rapidly progressive interstitial lung disease.^7^ We therefore anticipated that anti-Ro52 positivity might have a positive association with cardiac pathologies, which can be associated with inflammatory myopathies as well as pulmonary hypertension.

We did not find an association with cardiomyopathy, conduction abnormalities, myocarditis, acute coronary syndromes, or pulmonary hypertension. Of note, strict inclusion criteria based on 2017 ACR/EULAR classification criteria for idiopathic inflammatory myopathies led to the removal of many patients who received clinical diagnoses of anti-synthetase syndrome from their longitudinal rheumatologists, but did not have anti-Jo1 antibodies. As a result, many patients with amyopathic forms of anti-synthetase syndrome may have been excluded. Taken together, our observations suggest an unexpected disparate prognostic value of anti-Ro-52 antibodies in terms of risk of pulmonary and cardiac complications.

Pulmonary hypertension is typically associated with the development of right ventricular dysfunction and eventually right bundle branch block. The fact that in our cohort we see a possible trend toward protection from earlier-onset right bundle branch block was unexpected, and may reflect the small sample size for this rare disease, survivorship bias for patients who were able to establish at the study center, and/or removal of patients from the analysis who received a clinical diagnosis of anti-synthetase syndrome but were excluded based on strict adherence to the 2017 ACR/EULAR classification criteria for idiopathic inflammatory myopathies. Although anti-Ro-52 antibodies have a well-described association with pulmonary comorbidities, specifically interstitial lung disease and pulmonary hypertension, it appears their heterogeneous effects may result in a tension between increased secondary cardiac comorbidities due to pulmonary pathology and reduced primary cardiac comorbidities.

Our study has several strengths. To our knowledge, this is the first study to explicitly examine the association of anti-Ro-52 positivity with cardiac comorbidities in patients with anti-synthetase syndrome. We analyze a prospectively, well-characterized cohort in a highly specialized myositis center, resulting in a large cohort for this otherwise rare disease. In addition, we define pulmonary hypertension based on invasive hemodynamics (the gold standard for this diagnosis) and the integration of both frequency analyses and time-to-event analyses allows for a more granular description of the associations between anti-Ro-52 positivity and cardiac comorbidities and indirect internal validation of our findings.

Our study also has key limitations that are tightly connected to its strengths. Namely, its single center nature might limit its applicability to other cohorts and our limited sample size limits our statistical power. In addition, pulmonary hypertension was confirmed via invasive hemodynamics only in a subset of patients. Confirmation of pulmonary hypertension in a select group of patients may have also introduced a verification bias in our subgroup analysis. Due to survivorship bias and unclear duration of treatment of immunosuppression, our data do not account for timing and duration of immunosuppressive therapy. In addition, despite the relatively large size of our cohort for this rare condition, the number of cardiovascular events recorded was small, limiting the statistical power of our analyses. These limitations highlight the importance of increased clinician education regarding early identification of idiopathic inflammatory myopathies, anti-synthetase syndrome, and interstitial lung disease; further research into forms of classification that include patients with novel anti-synthetase antibodies; and routine comprehensive serotyping of patients with idiopathic inflammatory myopathies

The results of our study open multiple interesting avenues for further research. Understanding the biological basis of the disparate effect of other myositis-associated antibodies on lung and cardiac complications might highlight novel insights in the pathophysiology of the disease.

Future studies may also seek to further elucidate the details of the association between anti-Ro-52 positivity and pulmonary hypertension, including evaluation of pulmonary function tests to differentiate between group I and group III pulmonary hypertension. With the advent of accessible, load-independent metrics of right ventricular function, future studies may seek to evaluate the presence of occult right ventricular dysfunction in patients with anti-Ro-52 positivity.^19^ Finally, future studies may seek to evaluate if these effects are similarly seen across patients with Ro-52 positivity but alternative rheumatologic conditions.

## Data Availability

All data produced in the present study are available upon reasonable request to the authors

## Notes

### Competing Interest Statement

The authors have declared no competing interest.

### Author Declarations

The Institutional Review Board of Johns Hopkins Medicine gave ethical approval for this work

