## Supplementary figures and images for "Association of anti-Ro-52 positivity with cardiovascular outcomes in patients with anti-synthetase syndrome"

### Supplement 2

**Supplement 2.** Kaplan-Meier curves from time-to-event analyses of cardiovascular outcomes.

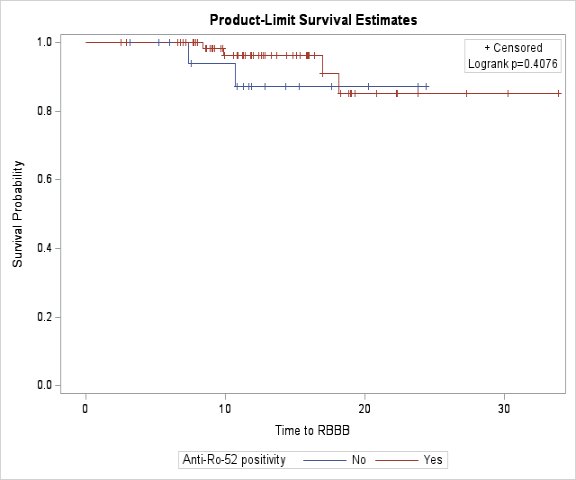

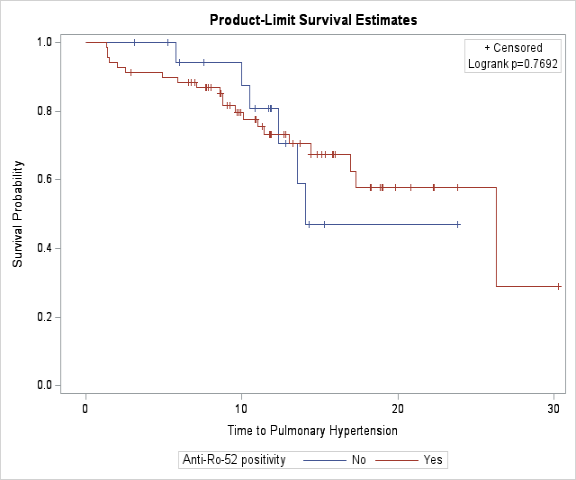

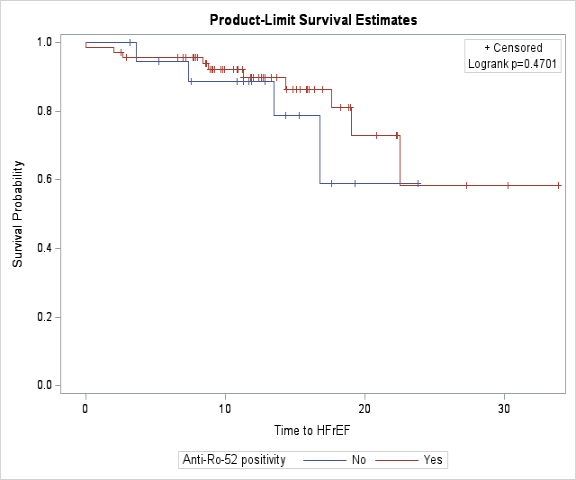

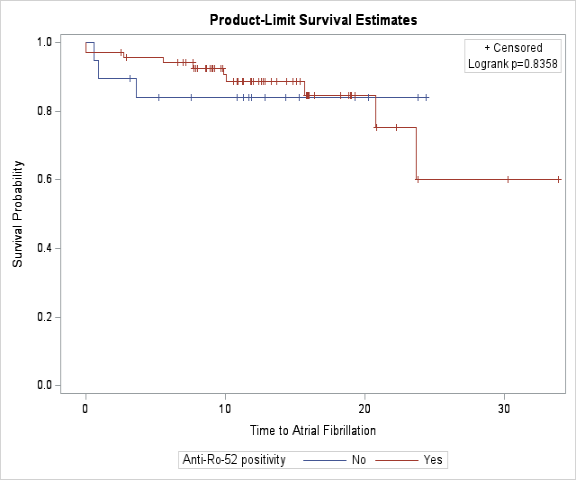

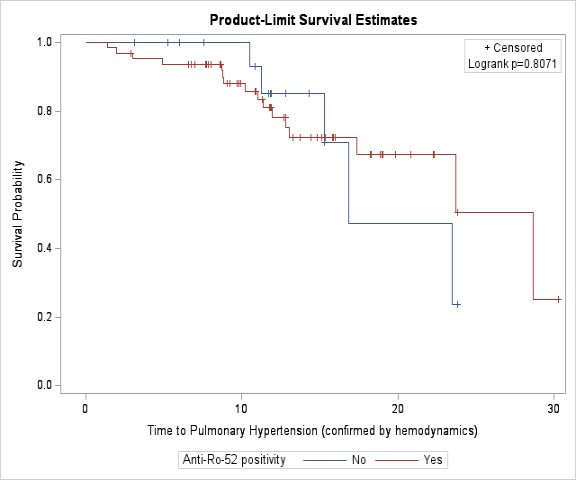
