## Supplement 1 for "Association of anti-Ro-52 positivity with cardiovascular outcomes in patients with anti-synthetase syndrome"

**Supplement 1 – Definitions of Cardiovascular Outcomes**

**Atrial fibrillation**

- Needs to have cardiologist-read EKG with confirmed atrial fibrillation

**Left bundle branch block**

- QRS > 120 ms with LBBB morphology
- Needs to have cardiologist-read EKG with confirmed LBBB

**Right bundle branch block**

- QRS > 120 ms with RBBB morphology
- Needs to have cardiologist-read EKG with confirmed RBBB

**Pulmonary hypertension**

- Needs to have transthoracic echo with RVSP > 35 mmHg
- For invasive hemodyanmic assessment, needs to have documented right heart cat with mPAP > 20 mmHg, PAWP < 15 mmHg, and PVR > 2 WU

**HFrEF**

- Needs to have TTE with EF less than or equal to 40% (TEE for emergent cardioversion and stress echo don’t count)
- Required descriptive variable: nadir EF

**ACS**

- Needs to either have had cath lab activation with LHC showing evidence of ACS (+/- stent) OR medical management of acute MI
- If diagnosis of type II myocardial infarction is given, it is NOT ACS

**Revascularization**

- Needs to have note documenting stent placement or coronary artery bypass grafting
- Required descriptive variables: free-text of each intervened-upon vessel, number of stents, CABG if completed with specified grafts, date of first ACS
- If revascularization was performed outside the context of ACS, should still document descriptive variables above and date of revascularization

**Myocarditis**

- Needs to have diagnosis of myocarditis in chart AND one of either (1) elevated troponin, (2) histopathologic diagnosis, (3) CMR read as consistent with myocarditis
- Required descriptive variable: form of diagnosis (if multiple present, prioritize pathology, then CMR, then troponin)
